# FAPα-positive fibroblasts in expert-reviewed colorectal hyperplastic polyps identify patients at increased risk of metachronous adenoma: a retrospective cohort study

**DOI:** 10.64898/2026.07.02.26357112

**Authors:** Nicolas Fenie, Julien Palasse, Marie-Bernadette Delisle, Audrey Ferrand

## Abstract

**Aims:** Serrated lesions contribute substantially to colorectal cancer (CRC), while routine management of small distal hyperplastic polyps (HPs) assumes low risk. Surveillance guidelines nevertheless incorporate uncertainty at the HP/SSL interface and recommend shortened intervals for large serrated lesions. (3,4) We tested whether fibroblast activation protein-α (FAPα) expression by stromal fibroblasts within expert-reviewed HPs stratifies risk of subsequent neoplasia.

**Methods and results:** In a single-centre historical cohort, FAPα immunohistochemistry (Abcam ab53066, 1:200) was performed on FFPE colon tissues from 64 patients (normal colon n=10; HP n=39; low-grade TA n=6; high-grade TA n=4; adenocarcinoma n=5). FAPα-positive stromal fibroblasts were quantified in 20 randomly selected fields at ×1000 by two blinded readers (ICC 0.93).

Among 39 patients with expert-reviewed index HPs and colonoscopic follow-up, the endpoint was metachronous adenoma occurring in the same general colonic area as the index HP, with proximal defined as ascending colon and distal as descending colon. Follow-up colonoscopies were scheduled every 2 years for up to 10 years. ROC analysis identified an optimal threshold of ≥9 FAPα-positive fibroblasts (AUC 0.8658; sensitivity 81.25%, specificity 87.93%).

FAPα-high status (44% of HPs) was associated with shortened neoplasm-free survival (log-rank p=0.0012): five-year neoplasm-free survival 41% versus 91% for FAPα no/low. In multivariable Cox modelling, FAPα-high status remained independently associated with metachronous adenoma (HR 4.5, 95% CI 1.2–16.8, p=0.022).

**Conclusion:** FAPα+ fibroblasts in expert-reviewed colorectal HPs identify a high-risk subgroup for metachronous adenoma, supporting stromal activation markers as a feasible pathology-anchored stratification tool.

## Introduction

Colorectal cancer (CRC) prevention is tightly linked to accurate identification and complete removal of precursor lesions at colonoscopy. Beyond conventional adenomas, serrated colorectal lesions are now recognised as major contributors to CRC, including a disproportionate share of interval cancers, reflecting both lesion biology and detection challenges (1). Within the serrated spectrum, hyperplastic polyps (HPs) are the most common lesions and, when small and distal, are generally regarded as low-risk findings in that small, distal HPs do not usually warrant shortened surveillance intervals (1).

A key source of uncertainty is the HP/SSL diagnostic boundary. The 2019 WHO criteria lowered the architectural threshold for SSL diagnosis (“a single unequivocally distorted crypt”), which increases sensitivity but also makes classification more dependent on sampling quality and observer judgement. In parallel, interobserver variability remains a documented limitation in distinguishing HP from SSL, even under simplified criteria, with direct consequences for surveillance decisions (2). Consequently, surveillance recommendations operationalise risk primarily through lesion size, multiplicity, location and dysplasia, while explicitly acknowledging diagnostic variability. For example, the US Multi-Society Task Force recommends colonoscopy in 3–5 years for HP ≥10 mm, favouring the shorter interval when there is concern regarding consistency in distinguishing SSL from HP, bowel preparation, or completeness of excision (3). European guidance similarly recommends 3-year surveillance after complete removal of any serrated polyp ≥10 mm or with dysplasia (4).

While such algorithms are pragmatic, they rely primarily on size, multiplicity, location and dysplasia, and they do not address whether a subset of lesions diagnosed as “HP” may carry biological risk signals beyond gross morphology. Nonetheless, biomarker-based evidence indicates that clinically meaningful heterogeneity exists within HPs: We previously reported that progastrin expression in HPs is associated with subsequent colorectal neoplasia, supporting the idea that common HPs may act as reporter lesions for patient-level susceptibility or a permissive mucosal context (5).

Most serrated biomarker work has focused on epithelial pathways. However, the microenvironment is recognised as a determinant of neoplastic evolution and clinical behaviour. Cancer-associated fibroblasts (CAFs) represent a major stromal component across epithelial cancers and can modulate extracellular matrix organisation, epithelial plasticity, and immune response (6,7). Within CAF-associated markers, fibroblast activation protein-α (FAPα), a membrane serine protease enriched in activated fibroblasts in epithelial malignancies, has established clinicopathological relevance in colon cancer (8). Importantly, FAP also has direct translational implication since FAPI-based theranostics and PET imaging are already advancing clinically, making FAP a practical bridge between pathology and scalable diagnostics (9).

Critically, there is still no reference framework that systematically characterises fibroblast activation states in pure HPs and benchmarks them against other serrated or adenomatous lesions.

Here, we test whether FAPα-positive stromal fibroblasts in expert-reviewed HPs stratify the risk of metachronous neoplasia.

## Materials and methods

### Study design

Single-centre historical cohort study.

### Study population and case selection

The required sample size for the primary endpoint, neoplasm-free survival (NFS), was calculated for a log-rank comparison using the Freedman–Schoenfeld method in STATA v11 (StataCorp). A total of 38 patients was required. Our study included colon tissue sections from 64 patients: 39 patients with colorectal hyperplastic polyps (HPs), 6 with low-grade tubular adenomas, 4 with high-grade tubular adenomas, 5 with adenocarcinomas, and 10 normal colonic tissues obtained from resections for non-complicated diverticular disease. The characteristic of the 39 patients presenting with HP have been previously published (5). Approval of an institutional research ethics committee was obtained in accordance with the precepts of the Helsinki Declaration.

We retrospectively reviewed the medical records of all patients with hyperplastic polyps diagnosed in the Department of Pathology at Rangueil Hospital (Toulouse, France) between 1 January 2000 and 31 December 2001. None had preceding colorectal adenocarcinoma or adenoma, familial colorectal adenocarcinoma history, serrated polyposis, chronic inflammatory bowel disease, insufficient colon site information or follow-up data, and none had evidence of colonic adenoma at the initial colonoscopy. Colonoscopy follow-up data were available for all patients included in the HP outcome analysis.

### Expert pathological re-review and diagnostic criteria

All index lesions included in the HP cohort were confirmed as hyperplastic polyps after expert pathological re-review performed by two gastrointestinal pathologists, blinded to clinical outcome and FAPα status. No additional levels were cut for re-review (assessment was performed on the available H&E material). Lesions showing features consistent with alternative serrated entities were excluded from the HP cohort.

### Outcome definition and surveillance schedule

The outcome event was metachronous colorectal adenoma occurring in the same general colonic area (proximal versus distal colon) as the index HP as determined from colonoscopy and pathology records, with proximal defined as ascending colon and distal defined as descending colon.

Follow-up colonoscopies were performed for surveillance at a standardised interval of 2 years, for up to 10 years, unless a metachronous event occurred earlier.

### Immunohistochemistry

Resected tissues were formaldehyde-fixed and paraffin embedded. Citrate buffer was used for epitope retrieval. The antibodies were applied overnight and detection was done using the DakoCytomation Envision Plus System-HRP. The anti-FAPα antibody (Abcam, ab53066) was used at a 1:200 dilution.

### Quantification and scoring reproducibility

FAPα expression was quantified as the number of FAPα-positive stromal fibroblasts counted in 20 randomly selected high-power fields per lesion at ×1000 magnification. Positive (CRC section) and negative (healthy/normal colon section) technical controls were included in each staining run. FAPα staining was scored independently by two observers in a double-blind fashion under pathologist supervision; Because inter-rater agreement was excellent (ICC = 0.93), values were reported as the average of the two readers. For patients with several polyps, the polyp with the widest hyperplastic area was retained for evaluation.

### Statistical analysis

To derive a binary classifier for risk stratification, an ROC analysis was performed using metachronous adenoma occurrence as endpoint; the optimal cut-off corresponded to ≥9 FAPα-positive fibroblasts, yielding sensitivity 81.25% and specificity 87.93% (AUC 0.8658). Neoplasm-free survival was evaluated using Kaplan-Meier estimates and log-rank tests. Cox proportional-hazards models were used to test the simultaneous influence of covariates with univariate p<0.20; backward stepwise selection retained variables with p<0.05. Analyses were performed using STATA v11; all tests were two-sided; statistical significance was set at p=0.05.

## Results

### FAPα-positive fibroblasts increase across normal colon, HPs, adenomas and adenocarcinoma

Representative pictures of FAPα staining in normal tissue, hyperplastic polyps (HP), adenoma and adenocarcinoma are shown in Figures 1 and 2.

**Figure 1.**
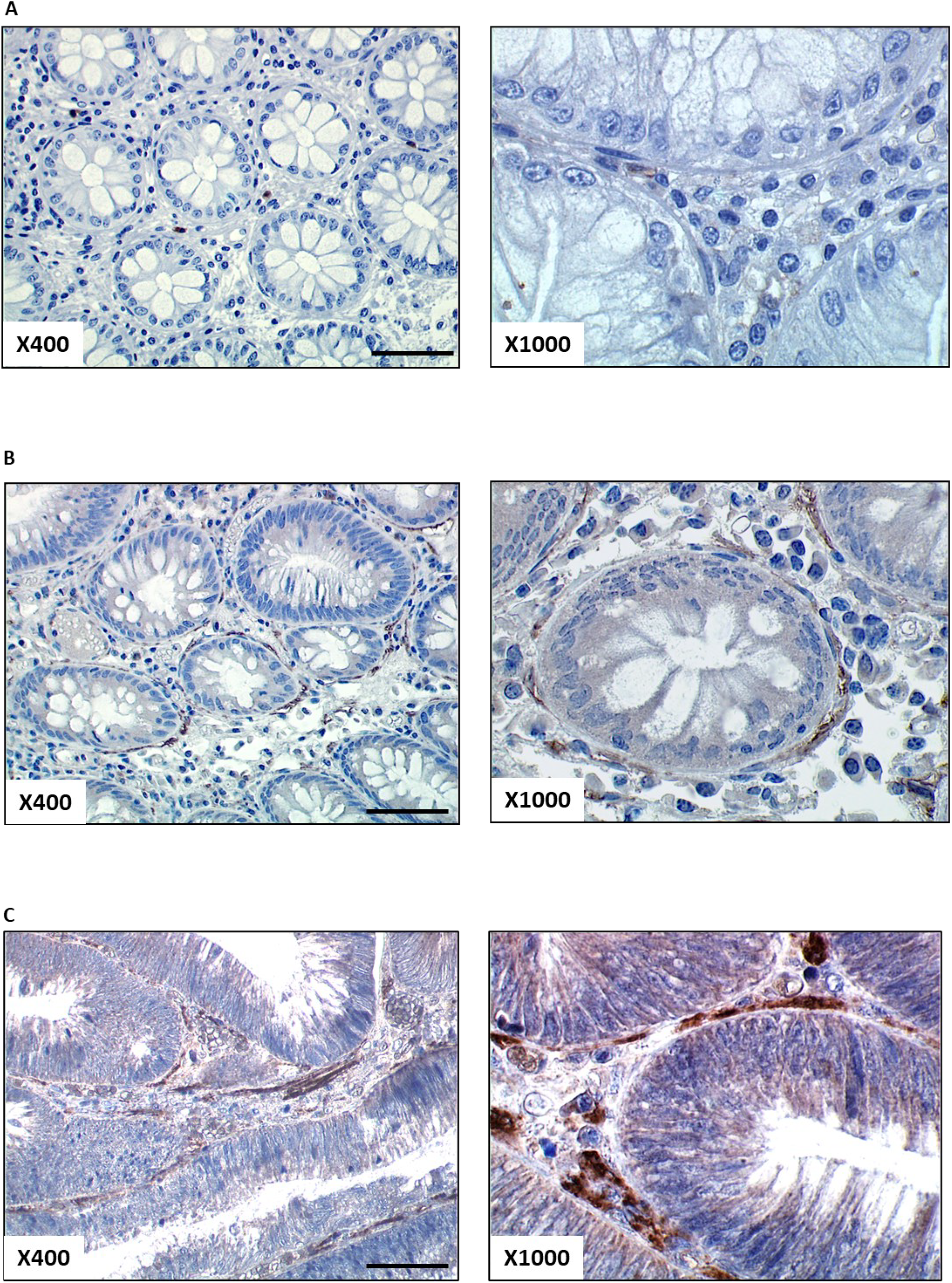
FAPα expression in normal colon, adenoma and adenocarcinoma tissues. Representative pictures for FAPα staining in normal colon (×400; ×1000), adenoma (×400; ×1000), and adenocarcinoma (×400; ×1000). Commercially available anti-FAPα antibody (Abcam ab53066) was used.

**Figure 2.**
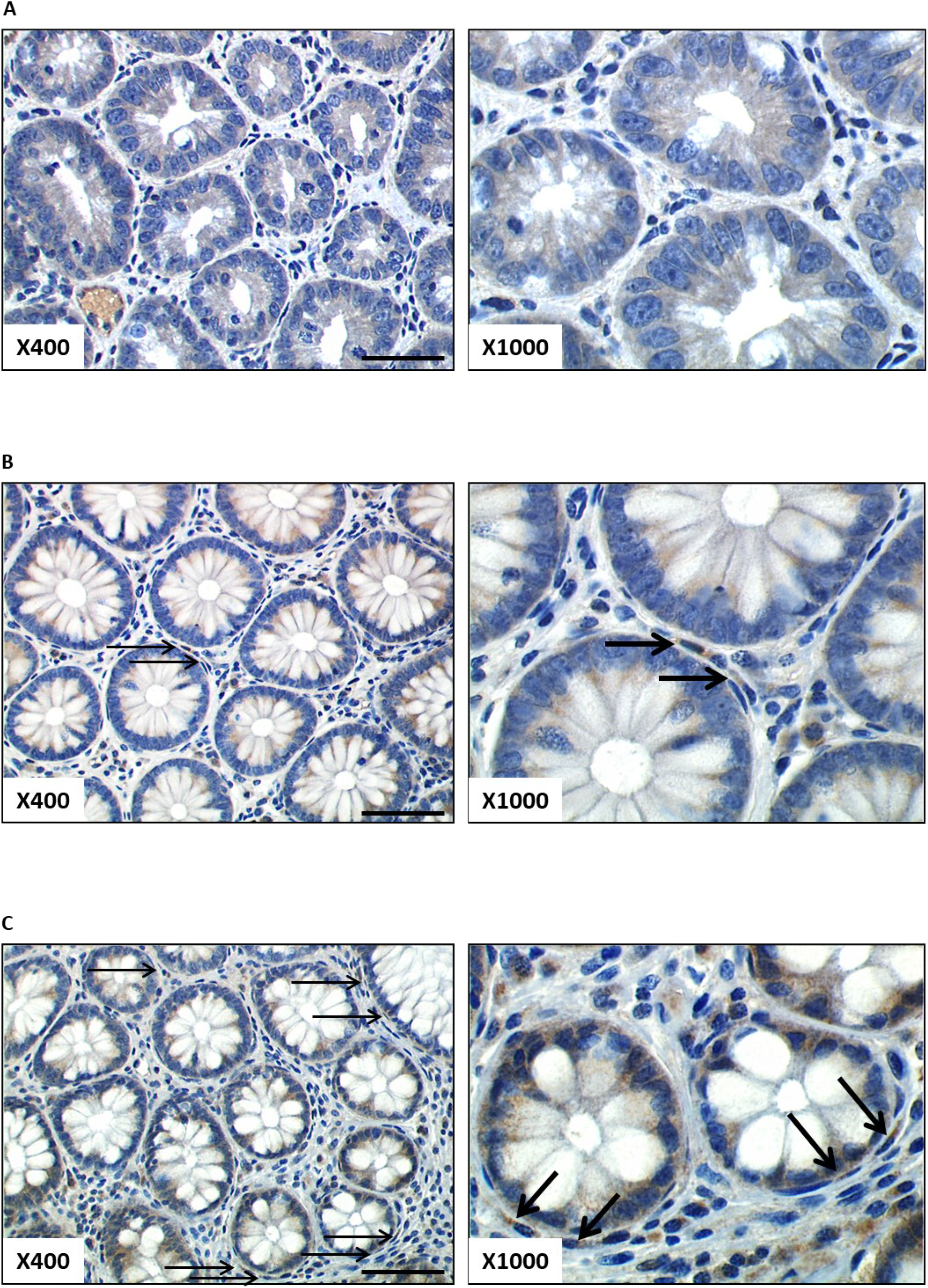
FAPα staining in hyperplastic polyps. Representative pictures for no/low/high FAPα staining in HP. Arrows indicate FAPα-stained fibroblasts.

The number of FAPα-positive fibroblasts in normal colon, HP, adenomas, and adenocarcinomas is reported in Figure 3. In the whole cohort, the number of FAPα-expressing fibroblasts was 10.33±1.394 (n=64). The number of FAPα-positive fibroblasts in normal colon from non-complicated diverticula (2.750±0.588, n=10) was significantly different from the number in HP (7.885±1.056, n=39, p=0.0195), low grade adenoma (11.92±1.044, n=6, p<0.0001), high grade adenoma (24.50±9.515, n=4, p=0.0025) and adenocarcinoma (31.30±7.782, n=5, p<0.0001). A significant difference was also found between HP and high-grade adenoma (p=0.0004) or adenocarcinoma (p<0.0001) while none was observed between HP and low-grade adenoma.

**Figure 3.**
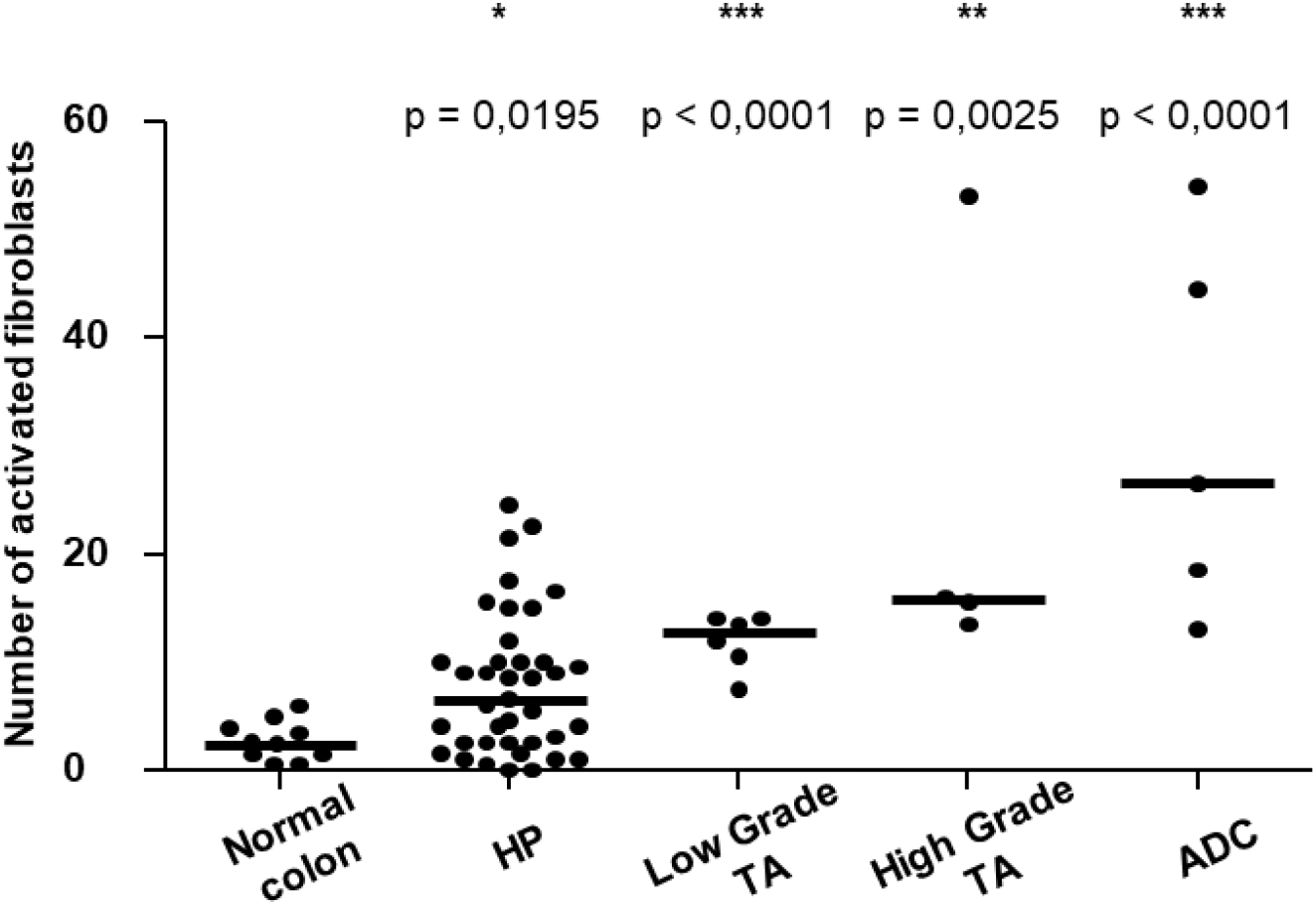
FAPα expression across lesion categories. Number of FAPα-positive fibroblasts in normal colon, hyperplastic polyps (HP), low-grade tubular adenoma (TA), high-grade tubular adenoma (TA), and adenocarcinoma (ADC).

### ROC analysis defines FAPα-high hyperplastic polyps

Surveillance colonoscopies were scheduled every 2 years for up to 10 years, providing a standardised follow-up framework across patients.

To assess whether FAPα predicts neoplasm occurrence after an index hyperplastic polyp, ROC analysis of the number of FAPα-positive fibroblasts yielded an AUC of 0.8658 (95% CI, 0.75–0.98). The threshold between ‘no/low’ and ‘high’ FAPα expression in HP was determined to be 9 stained fibroblasts to obtain the optimal specificity–sensitivity couple (sensitivity 81.25% and specificity 87.93%) (Figure 4). Thus, FAPα staining corresponded to no/low expression when the number of stained fibroblasts was <9 and high expression when ≥9. In this cohort, 56% (95% CI: 39–72) had no/low FAPα expression while 44% (95% CI: 27–60) had high FAPα expression (Table 1).

**Table 1.** Clinical and immunohistological features. FAP expression was recorded as no/low or high based on the number of stained fibroblasts counted in 20 randomly picked fields at ×1000 magnification. The threshold in HP was 9 stained fibroblasts based on ROC analysis. Follow-up colonoscopies were scheduled every 2 years for up to 10 years.

| Table 1. Clinical and immunohistological features |  |
| --- | --- |
| Variables | Patients with coloscopic follow up<br>(ref Do et al) |
|  | n=39 |
|  | percentage [95%CI] |
| **Age (y.o)** |  |
| Median | 64 |
| Range | 33-89 |
| **Sex** |  |
| Male | 59% [42-74] |
| Female | 41% [26-58] |
| **Number of polyps** |  |
| 1 | 62% [45-77] |
| 2-4 | 38% [23-55] |
| **Size of polyps (largest diameter, mm)** |  |
| Median | 3 |
| Range | 1-5 |
| **Localisation** |  |
| Proximal colon | 18% [8-34] |
| Distal colon | 82% [66-82] |
| **Occurrence of metachronous adenoma** | 41% [26-59] |
| **Expression of FAP** |  |
| Low / No expression (<9 stained fibroblasts) | 56% [39-72] |
| High expression ($\geq 9$ stained fibroblasts) | 44% [27-60] |

**NOTE:** FAPα expression was recorded as “No/low expression” or “high expression” based on the number of stained fibroblasts counted in 20 randomly picked microscope fields at X1000 magnification. The threshold of FAPα expression in HP was determined to be 9 stained fibroblasts, 9 corresponding to the optimal specificity/sensibility couple based on the ROC analysis,
95% CI: Bionomial exact 95% confidence index was calculated for each percentage.

**Figure 4.**
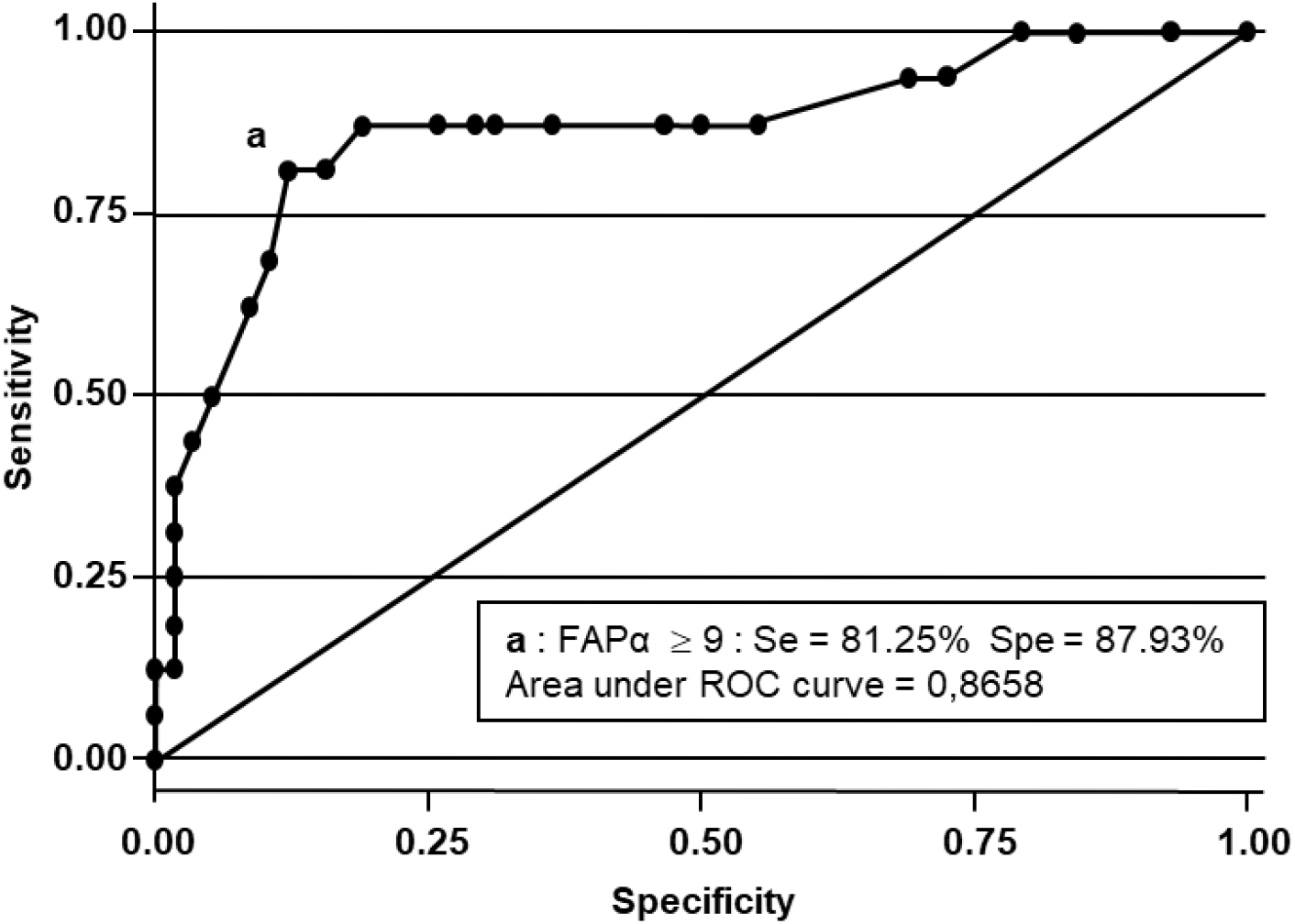
ROC curve for prediction of metachronous adenoma. ROC curve for FAPα staining in HP. Threshold ‘a’ corresponds to ≥9 FAPα-stained fibroblasts, with sensitivity 81.25% and specificity 87.93%; AUC 0.8658.

### FAPα-high status is associated with shortened neoplasm-free survival

Only occurrence of metachronous adenoma in the same general colonic area (proximal corresponds to ascending colon; distal corresponds to descending colon) was considered as a recurrence event.

Among patients with expert-reviewed HPs (n=39) and colonoscopy follow-up, 41% (95% CI: 26-59) developed a metachronous adenoma in the same general colonic area (proximal vs distal) (Table 1).

Kaplan–Meier analysis demonstrated significantly shorter neoplasm-free survival for the FAPα-high group (log-rank p=0.0012) (Figure 5). Five-year neoplasm-free survival was 91% (95% CI 68-98) for no/low FAPα versus 41% (95% CI 19-63) for FAPα-high HPs. Patients with high FAPα expression had a median neoplasm-free survival of 5 years, whereas during the 10-year follow-up, the median survival was not reached in patients with no/low FAPα expression (Table 2; Figure 5).

**Table 2.** Neoplasm-free survival according to clinical and immunohistological features. Multivariate analysis was conducted using a Cox model including all variables with a p value <0.20 in univariate analysis. Initial model included age, size, localisation and FAP expression; final model retained age and FAP. Follow-up colonoscopies were scheduled every 2 years for up to 10 years.

95% CI: Bionomial exact 95% confidence index was calculated for each percentage.
| Table 2. Neoplasm-free survival according to clinical and immunohistological features |  |  |  |  |  |
| --- | --- | --- | --- | --- | --- |
| Variables | Neoplasm-free Survival |  |  |  |  |
|  | Univariate analysis |  |  | Multivariate analysis * |  |
|  | Median (y) | 5 y.S - [95% CI] | p-value | Hazard Ratio (HR) [95%CI] | p-value |
| **Age (y.o)** |  |  | <0.0001 |  | 0.001 |
| <64 y.o | 10 | 95% [72-99] |  | - |  |
| ≥64 y.o. | 5 | 26% [7-51] |  | 14.5 [12.9-72.5] |  |
| **Sex** |  |  | 0.4658 # |  |  |
| Male | NR | 77% [54-90] |  |  |  |
| Female | 10 | 51% [23-73] |  |  |  |
| **Number of polyps** |  |  | 0.4064 |  |  |
| 1 | 10 | 74% [51-87] |  |  |  |
| 2-4 | 7 | 56% [27-78] |  |  |  |
| **Size of polyps (largest diameter, mm)** |  |  | 0.0637 # |  | 0.895 # |
| <3mm | NR | 76% [47-90] |  | - |  |
| 3-6mm | 7 | 60% [35-78] |  | 1.1 [0.4-3.2] # |  |
| **Localisation** |  |  | 0.1378 # |  | 0.660 # |
| Proximal colon | 5 | 43% [10-73] |  | - |  |
| Distal colon | 10 | 74% [54-86] |  | 1.3 [0.4-4.1] # |  |
| **Expression of FAP** |  |  | 0.0012 |  | 0.022 |
| Low/no expression (<9) | NR | 91% [68-98] |  | - |  |
| high expression (≥9) | 5 | 41% [19-63] |  | 4.5 [1.2-16.8] |  |

**Note: Neoplasm-Free Survival among patients with colonic hyperplastic polyps who had a colonoscopic follow-up. Multivariate analysis was conducted using a Cox model including all variables with a P value <0.20 in the univariate analysis.**
\* The initial Cox model included the age, the size, the localisation of the polyps and FAPα expression. Final model was obtained after backward stepwise selection, keeping only the variables with a P value <0.05 (i.e. age and FAPα ). 95% CI was calculated for all 5-year survival values.

**Figure 5.**
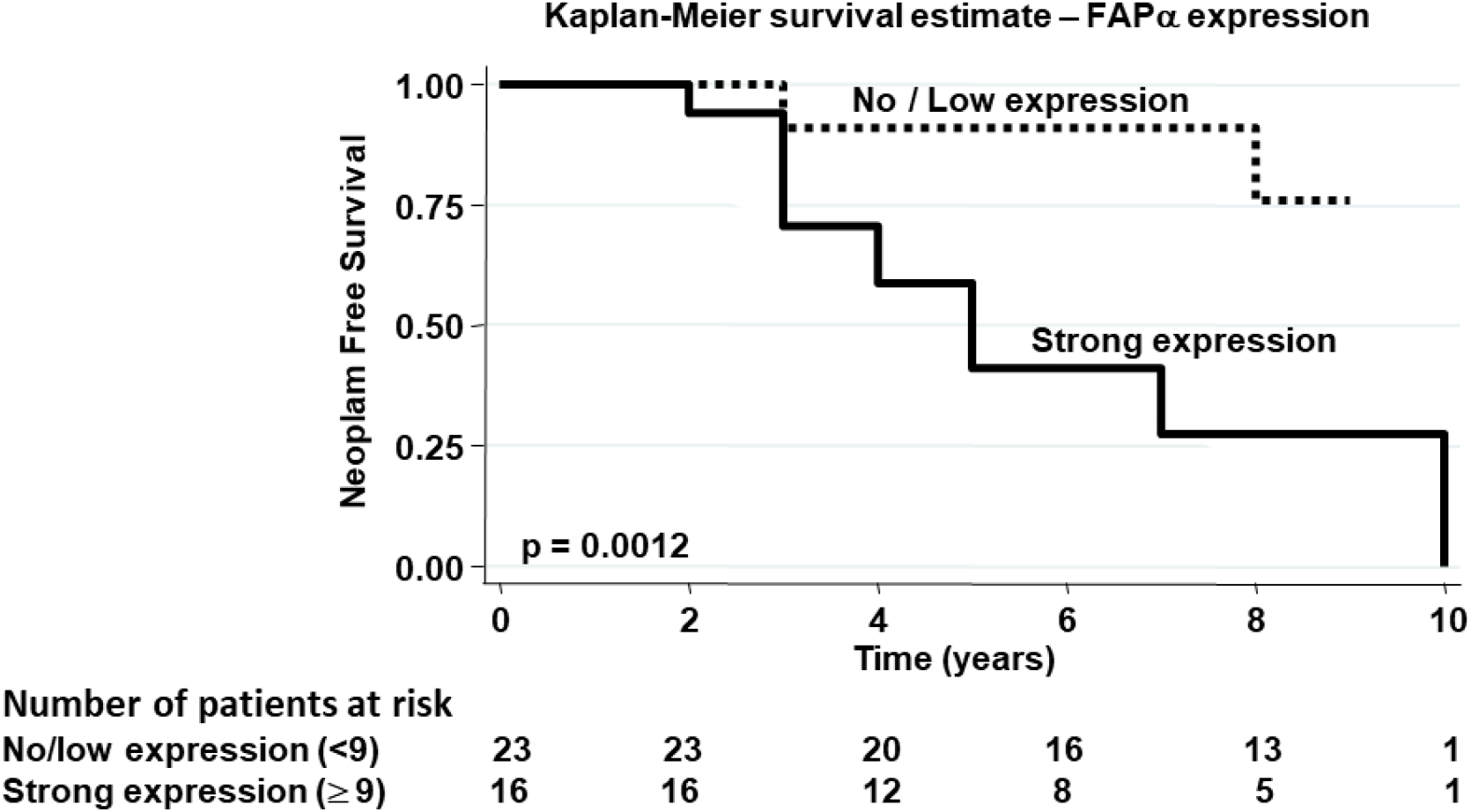
Kaplan-Meier neoplasm-free survival estimate based on FAPα expression in HP patients. Neoplasm-free survival among patients with colonoscopic follow-up according to FAPα expression. Endpoint: metachronous adenoma in the same general colonic area (proximal = ascending colon; distal = descending colon). p-value corresponds to log-rank analysis.

### Multivariable Cox modelling

In multivariable Cox regression, FAPα-high status remained independently associated with metachronous adenoma (HR 4.5, 95% CI 1.2–16.8, p=0.022), alongside age in the final model (Table 2).

### Reporting and biomarker standards

The study is reported in line with REMARK principles for tumour marker prognostic studies: clear endpoint definition, blinded biomarker assessment, prespecified modelling strategy, and transparent reporting of effect sizes with confidence intervals. Internal and external validation are recognised as necessary steps prior to clinical translation.

## Discussion

This study demonstrates that FAPα-positive stromal fibroblasts are detectable in a substantial subset of expert-reviewed colorectal hyperplastic polyps (HPs) and that this phenotype identifies patients at increased risk of metachronous adenoma in the same general colonic area (proximal vs distal) during follow-up. Using quantitative immunohistochemistry and ROC-derived thresholding (≥9 FAPα+ fibroblasts across 20 ×1000 fields), FAPα-high HPs exhibited markedly reduced neoplasm-free survival and retained an independent association with outcome in multivariable modelling.

Recent reviews of serrated colorectal tumorigenesis emphasise that serrated trajectories are biologically distinct and clinically consequential but heterogeneous, spanning from early initiation events, progressive epigenetic remodelling to variable progression kinetics influenced by local tissue constraints and microenvironmental context (10). In this frame, our data do not argue that HPs are obligate cancer precursors. Rather, they support a “reporter lesion” model in which a subset of lesions classified as HP, after expert re-review, captures a local microenvironmental activation state permissive for subsequent neoplasia, consistent with the idea that early carcinogenesis is not purely epithelial (10). This is biologically plausible given the established role of activated fibroblasts in shaping extracellular matrix organisation, paracrine signalling, and immune contexture, and aligns with the broader concept that early carcinogenesis is not purely epithelial.

Indeed, CRC microenvironment literature increasingly supports the view that CAFs are heterogeneous and can regulate epithelial fitness, extracellular matrix organisation and immune response, thereby influencing tumour initiation/progression and therapeutic response (6,7,11). In this framework, FAPα is particularly attractive because it is a canonical CAF-associated marker with clinicopathological relevance in colon cancer (8). Meta-analyses across solid tumours also support associations between high FAP expression and adverse outcomes, consistent with FAP marking stromal programs linked to aggressiveness (12,13). Although most of the outcome evidence derives from invasive disease, these data strengthen the plausibility that detecting FAPα-positive fibroblasts in an index lesion classified as HP after expert review reflects a microenvironmental activation state relevant to neoplasia risk. CAF activation can encode early neolasia permissiveness.

A key mechanistic question is whether the stromal activation captured by FAPα in HPs is serrated-specific or represents a broader pro-neoplastic program. Notably, recent analysis suggests TGFβ-responsive stromal activation can occur early in serrated carcinogenesis, including BRAF-associated contexts (15). This provides a coherent biological bridge between serrated epithelial initiation and early stromal activation states and motivates follow-up studies integrating epithelial trajectory markers (e.g., BRAF/KRAS, CIMP/MMR where appropriate) with stromal programs (TGFβ/ECM/immune exclusion) in the same lesions.

FAPα has exceptional translational traction because FAP inhibitor (FAPI)-based imaging and theranostic strategies are already in clinical development and use, making FAPα a practical bridge between histopathology and scalable diagnostics (9). CRC-focused work further shows that FAPα expression can identify stromal-rich, poor-prognosis molecular phenotypes (e.g., CMS4) and can be detected by ^68^Ga-FAPI-PET/CT, supporting feasibility of FAP-centered stratification (14). In pathology workflows, a robust single-marker IHC readout could be a pragmatic adjunct to morphology-driven surveillance decisions, particularly in contexts where guidelines already acknowledge uncertainty (3,4).

A major strength is that index lesions were confirmed as HP after expert pathological re-review by two gastrointestinal pathologists blinded to outcome, reducing the likelihood that the signal is simply diagnostic drift at the HP/SSL boundary. Additional strengths include blinded dual quantitative scoring (with excellent inter-rater agreement (ICC 0.93), supporting measurement reproducibility), explicit ROC-derived thresholding, and time-to-event analyses demonstrating an independent association of FAPα-high status with outcome.

Limitations remain. First, the study is retrospective and single-centre, and the ROC-derived cut-off was selected within the same dataset used for outcome association, raising the possibility of optimistic discrimination estimates; independent validation (and ideally internal resampling such as bootstrap/cross-validation) is therefore required prior to clinical translation. Second, re-review did not include additional levels; while expert review reduces misclassification risk, the absence of deeper levels cannot fully exclude sampling-related diagnostic uncertainty in borderline serrated lesions. Third, the endpoint was metachronous adenoma rather than advanced serrated lesions or CRC, limiting inference on serrated-specific malignant progression. Finally, although follow-up was conducted on a standardised 2-year schedule for up to 10 years, residual detection bias related to colonoscopy quality (preparation quality, withdrawal time, serrated detection rate) cannot be fully excluded without endoscopy-quality metrics.

Given where the field is moving, the logical next step is spatially resolved and/or single-cell profiling to define CAF states, their neighbourhood relationships and their coupling to epithelial programs. Recent spatial and single-cell CRC studies provide methodological templates for extending the present observation to mechanism (16,17).

Overall, these findings support an up-to-date conceptual shift: microenvironmental activation states, measurable by routine pathology tools, may contribute to risk stratification even within lesions traditionally managed as low risk. In HPs, high prevalence and surveillance decisions often driven by coarse surrogates, FAPα-positive fibroblasts emerge as a plausible, actionable biomarker candidate warranting external validation and mechanistically integrated positioning.

## Data Availability

All data produced in the present study are available upon reasonable request to the authors

## Abbreviations

ADC: adenocarcinoma
CAF: cancer-associated fibroblast
CI: confidence interval
CRC: colorectal cancer
FAPα: fibroblast activation protein-α
HP: hyperplastic polyp
IHC: immunohistochemistry
ROC: receiver operating characteristic
SSL: sessile serrated lesion
TA: tubular adenoma
USMSTF: U.S. Multi-Society Task Force.

## Ethics approval

This cohort is part of the one we used in (5). Institutional research ethics committee approval; conducted according to the Declaration of Helsinki.

## Conflict of interest disclosure

No potential conflicts of interests were disclosed.

## Data availability statement

The data that support the findings of this study are available from the corresponding author upon reasonable request.

## Reporting support

ChatGPT (OpenAI) was used solely for grammatical and stylistic editing of manuscript text; it was not used for data analysis, figure generation, or interpretation of results.

## Notes

**Supported by:** This work was supported by grants from INSERM, Association pour la Recherche contre le Cancer (ARC N° A09/4/5033, ARC SFI 20111203828) and Plan Cancer ‘Biology des systèmes’ (Mocassin 2017, grant number C18006BS)

### Competing Interest Statement

The authors have declared no competing interest.

### Author Declarations

Approval of the institutional research ethics committee (comite de protection des personnes sud-ouest et outre-mer I) was obtained in accordance with the precepts of the Helsinki Declaration.

